# Epidemiological findings and estimates of the instantaneous reproduction number and control strategies of the first Mpox outbreak in Latin America

**DOI:** 10.1101/2023.09.15.23295629

**Authors:** Cándida Díaz-Brochero, Zulma M. Cucunubá

## Abstract

**Background and objective:** since 2022, the world has experienced the largest Mpox outbreak in history. The region of the Americas has been especially affected, accounting for 67% of the cumulative cases worldwide. Key epidemiological characteristics of the outbreak for the Latin American region remain understudied. Here we provide estimations of R(t) and describe the key epidemiological trends, and vaccination strategies of the Mpox outbreak in six Latin American countries.

**Materials and methods:** we investigated the public health response to the Mpox outbreak in six Latin American countries from official sources. The surveillance data were obtained from the official Mpox report of the Pan American Health Organization. We calculated cumulative and incident confirmed cases according to the report date for each country and represented these findings in epidemic curves. The R(t) was estimated on weekly sliding windows for each country.

**Results:** The maximum means of R(t) ranged from 2.28 to 3.15 from May to July 2022. At epidemiological week 42, the R(t) estimates were as follows: Argentina: 0.94 (95% Credible interval -CrI-0.77 to 1.12), Chile: 0.83 (95% CrI: 0.64 to 1.05), Colombia: 0.60 (95% CrI: 0.52 to 0.69), Mexico: 0.75 (95% CrI: 0.67 to 0.84). For Peru and Brazil, R(t) decreased to less than 1 in weeks 32 and 33, with estimates between 0.89 (95% CrI: 0.82 to 0.96) and 0.94 (95% CrI: 0.89 to 0.98), respectively.

**Conclusion:** our results provide relevant information about current trends and future scenarios of the Mpox outbreak in Latin America. From late August to early September 2022, R(t) started to decrease to values less than 1, despite the limited delivery of vaccination programs implemented across the region. However, a large population remains at risk, and there is a possibility of new waves of the disease as the epidemic continues its course.

## Introduction

Since January 2022, the world has experienced the largest mpox outbreak in history, with more than 89 thousand confirmed cases in 114 countries from January to the third week of August 2023. The World Health Organization (WHO) assessed the risk in the Region of the Americas as moderate, accounting for approximately 67% of the cumulative confirmed cases and 80% of total deaths. During 2023, the number of cases reported weekly has declined substantially from the global peak of 7,576 cases observed in the week of 08 Aug 2022. However, cases continued to be reported in the region in a lesser extent (1, 2)

The IHR Emergency Committee on the multi-country outbreak of mpox held its fifth meeting on 10 May 2023. Having considered the views of committee members and advisors, the WHO Director-General determined that this outbreak no longer constitutes a public health emergency of international concern and issued revised temporary recommendations for a transitional period towards a long-term mpox control strategy. One of the recommendations was the need to maintain epidemiological surveillance of mpox and to implement a coordinated research agenda to generate and disseminate evidence for key scientific and public health aspects of mpox prevention and control (3). In this way, it highlights the need to collect and analyze key epidemiological parameters in different regions, which can potentially serve as input for the development of mathematical models adjusted to local realities.

One epidemiological parameter that allows the approximation of transmissibility is the instantaneous reproductive number or R(t); this parameter is related to the speed of spread of an emerging infection and the risk of longer-term endemicity. It is defined as the number of secondary infections from a primary case in a partially susceptible population (4). Historically, the estimation of R(t) has been key in the planning of containment strategies and local and regional public health measures, being especially useful for assessing the trajectory of an epidemic and the impact of prioritizing control interventions (5, 6).

Here we provide estimations of R(t) and describe the early epidemiological trends of the (2022) Mpox outbreak in the six Latin American countries with the highest number of reported cases, which are: Argentina (ARG), Brazil (BRA), Chile (CHL), Colombia (COL), Mexico (MEX) and Peru (PER).

## Materials and methods

The surveillance data were obtained from the official Mpox report of the Pan American Health Organization (PAHO) (7) and in some cases from the official report of individualized cases for each country (8).

Descriptive analyses of the most relevant time, place and person variables were performed. We calculated the cumulative and incident confirmed cases according to the report date for each selected country, and represented these findings in epidemic curves, using the package “incidence” of the R programming language (9) through the Rstudio interface version 4.2.1 (10).

Then, the R(t) was estimated for each country by using the “estimate_R′′ function of the “EpiEstim” package (11), according to the methodology of Cori et al. (2017) (12).

We used a parametric distribution, by means of the “parametric_si” configuration of the package. For the estimation of the R(t) it was necessary to consider the serial interval (SI), which denotes the time from illness onset in the primary case to illness onset in the secondary case. Since there is no local SI data from Latin American countries, we used the values estimated by Madewell et al. 2022 (13) in 12 U.S. jurisdictions, from May to August 2022. Taking the above into account, we considered a mean SI of 8.5 days and standard deviation of 5.0 days, reported by the aforementioned study.

The R(t) was estimated on weekly sliding windows, given by the parameters “t_start” and “t_end”. The date of onset was used as the estimation date, with a lag of 14 days. The analysis was carried out using the R programming language through its Rstudio interface (version 4.2.1) (10).

A sensitivity analysis was conducted to allow for changes in the mean prior of the reproduction number between 1.5 and 3.8 as reported in the literature (14).

## Results

### Descriptive analysis of time, place and person variables

The top six countries in Latin America with the highest number of cases as of August 22, 2023, are in order: Brazil (n=10,967); Colombia (n=4,090); Mexico (n=4,050); Peru (n=3,812); Chile (n=1,442) and Argentina (n=1,130). Cumulative and new confirmed cases of Mpox discriminated by selected countries are represented in **figure 1** and a map of the cumulative Mpox confirmed cases is presented in **figure 2**.

**Figure 1:**
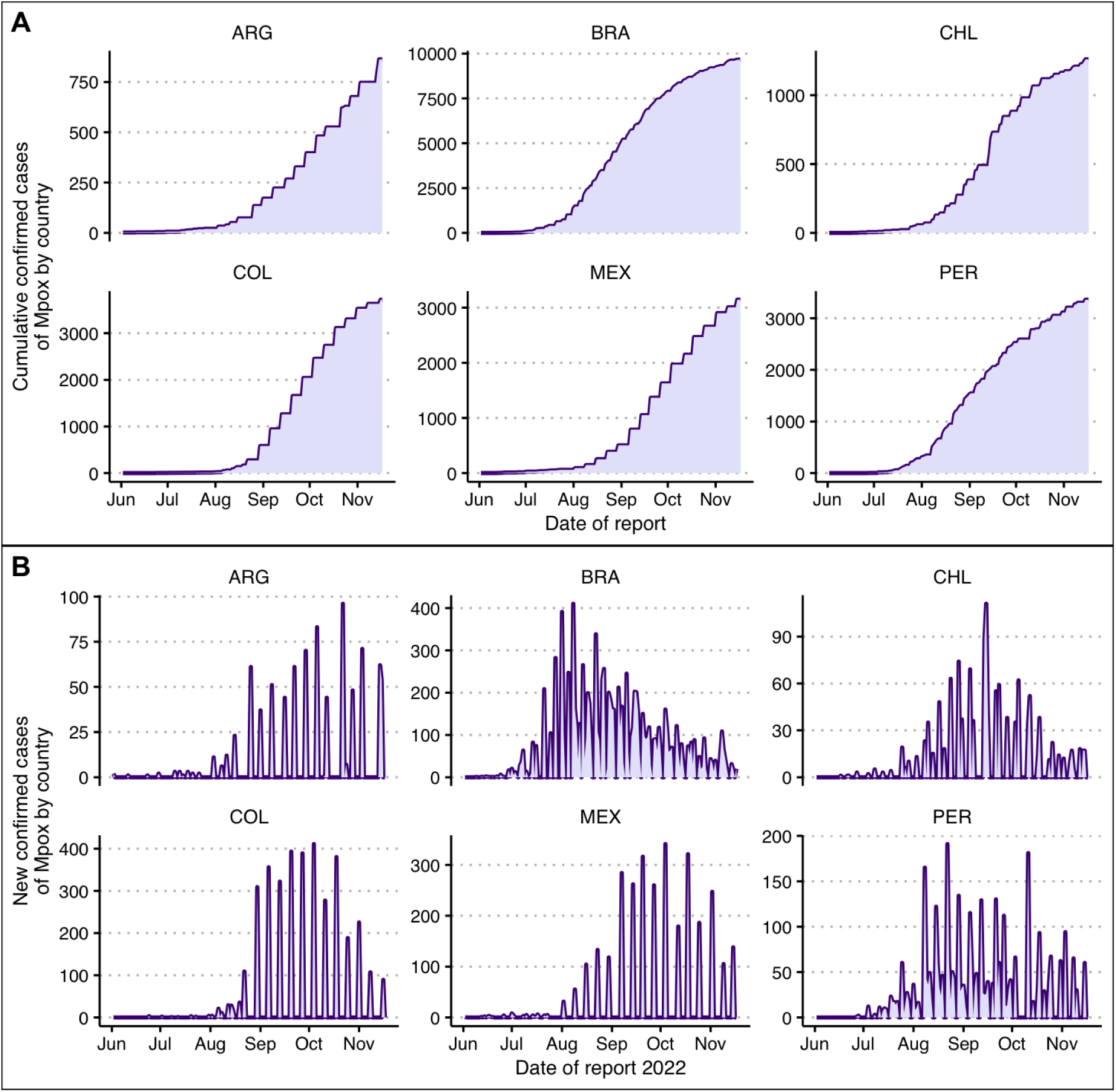
**A:** Cumulative confirmed cases of Mpox by country during 2022; **B:** New confirmed cases of Mpox by country during 2022

**Figure 2:**
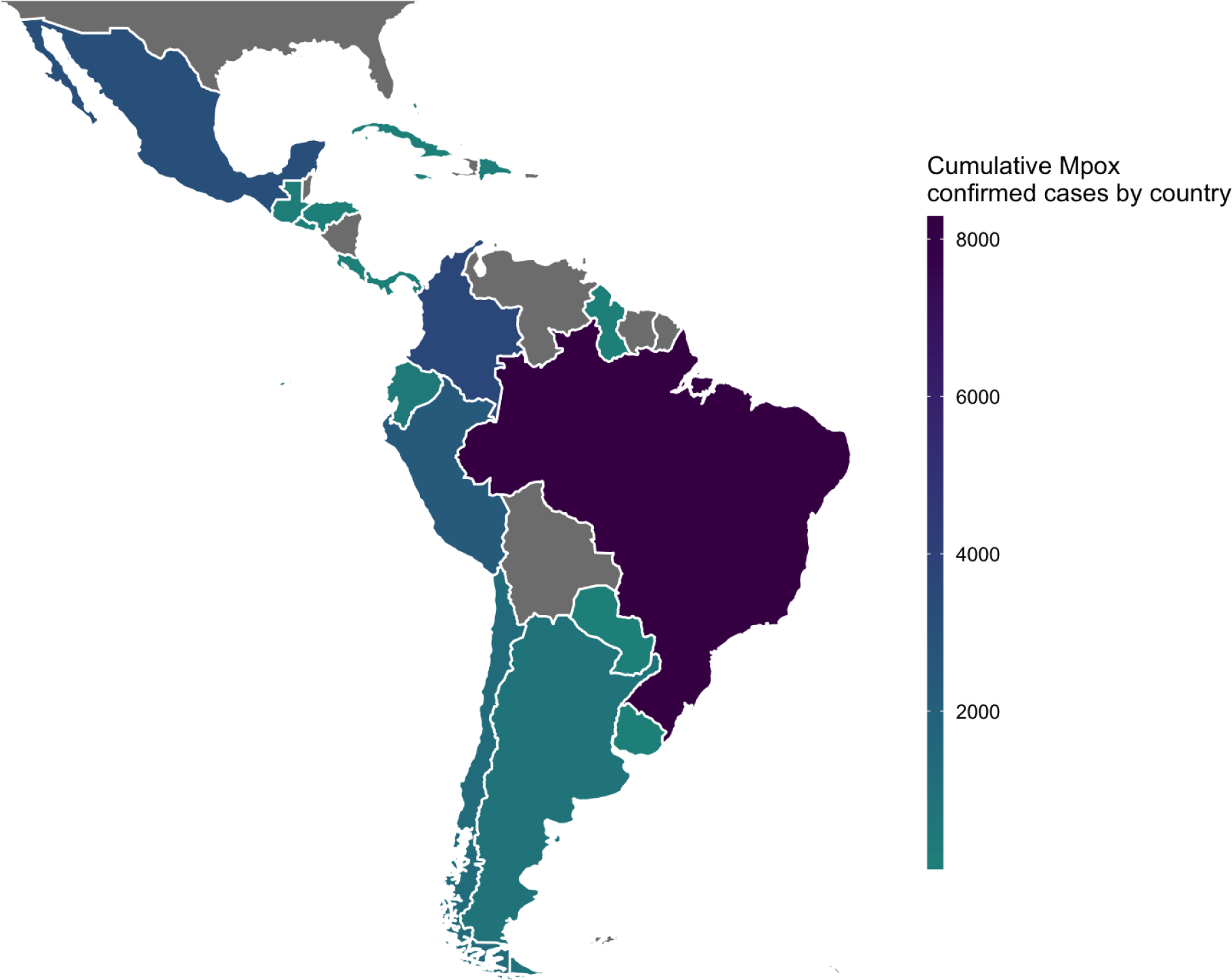
Map of the cumulative monkeypox confirmed cases from ARG, BRA, CHL, COL, MEX and PER

The main characteristics of the confirmed cases by country are presented in table 1.

**Table 1:** Main characteristics of the confirmed cases by country as 22 August 2023.

| Country | Proportion of male % | Median age in years | Number of deaths related to Mpox | Confirmed cases per million inhabitants |
| --- | --- | --- | --- | --- |
| ARG | 98.5 | 34 | 2 | 24.7 |
| BRA | 91.8 | 32 | 16 | 51.2 |
| CHL | 98.6 | 34 | 3 | 74.3 |
| COL | 97.1 | 31 | 0 | 79.4 |
| MEX | 97.5 | 33 | 30 | 31.9 |
| PER | 98.2 | 32 | 20 | 113.1 |

The proportion of males between the confirmed cases varies between 91.8% and 98.5%, with a median age that ranges between 31 and 34 years. The country with the greatest number of deaths is Mexico with 30 confirmed deaths secondary to Mpox, followed by Peru with 20. Regarding the number of cases per million inhabitants, the leading country is Peru with 113.1 mpox cases per million inhabitants, and the second is Colombia, with 79.4.

### Early instantaneous reproductive number R(t) estimations for each country

The maximum means of the instantaneous reproductive number R(t) as of November 18, 2022, ranged from 2.28 to 3.15 from May to July 2022, being higher in Colombia and Brazil, and lower in Mexico and Argentina (See **table 2**).

**Table 2:**
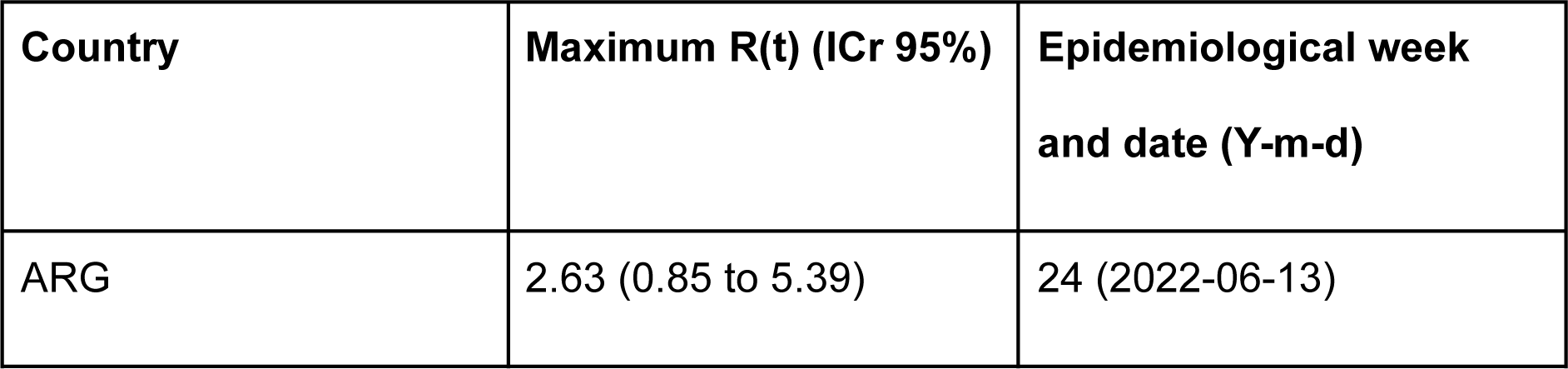
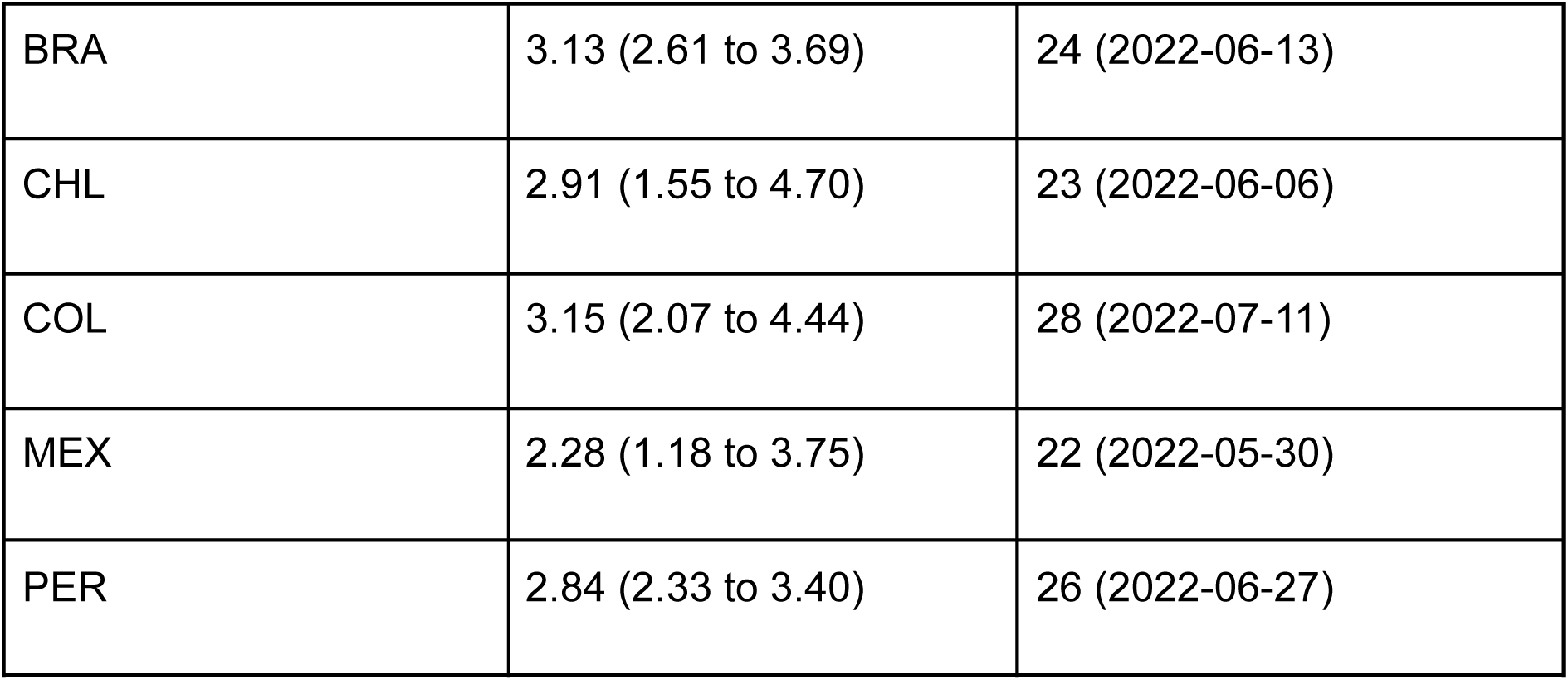
Maximum instantaneous reproductive number R(t) estimations for each country.

| Country | Maximum $R(t)$ (ICr 95%) | Epidemiological week and date (Y-m-d) |
| --- | --- | --- |
| ARG | 2.63 (0.85 to 5.39) | 24 (2022-06-13) |
| BRA | 3.13 (2.61 to 3.69) | 24 (2022-06-13) |
| CHL | 2.91 (1.55 to 4.70) | 23 (2022-06-06) |
| COL | 3.15 (2.07 to 4.44) | 28 (2022-07-11) |
| MEX | 2.28 (1.18 to 3.75) | 22 (2022-05-30) |
| PER | 2.84 (2.33 to 3.40) | 26 (2022-06-27) |

At epidemiological week 42 of 2022 -late October-the R(t) values were as follows: Argentina: 0.94 (95% Credible interval -CrI-0.77 to 1.12), Chile: 0.83 (95% CrI: 0.64 to 1.05), Colombia: 0.60 (95% CrI: 0.52 to 0.69), Mexico: 0.75 (95% CrI: 0.67 to 0.84). For Peru and Brazil, R(t) values decreased to less than 1 in epidemiological weeks 32 and 33 -late August 2022-with estimates between 0.89 (95% CrI: 0.82 to 0.96) for Peru and 0.94 (95% CrI: 0.89 to 0.98) for Brazil (See figure 3).

**Figure 3:**
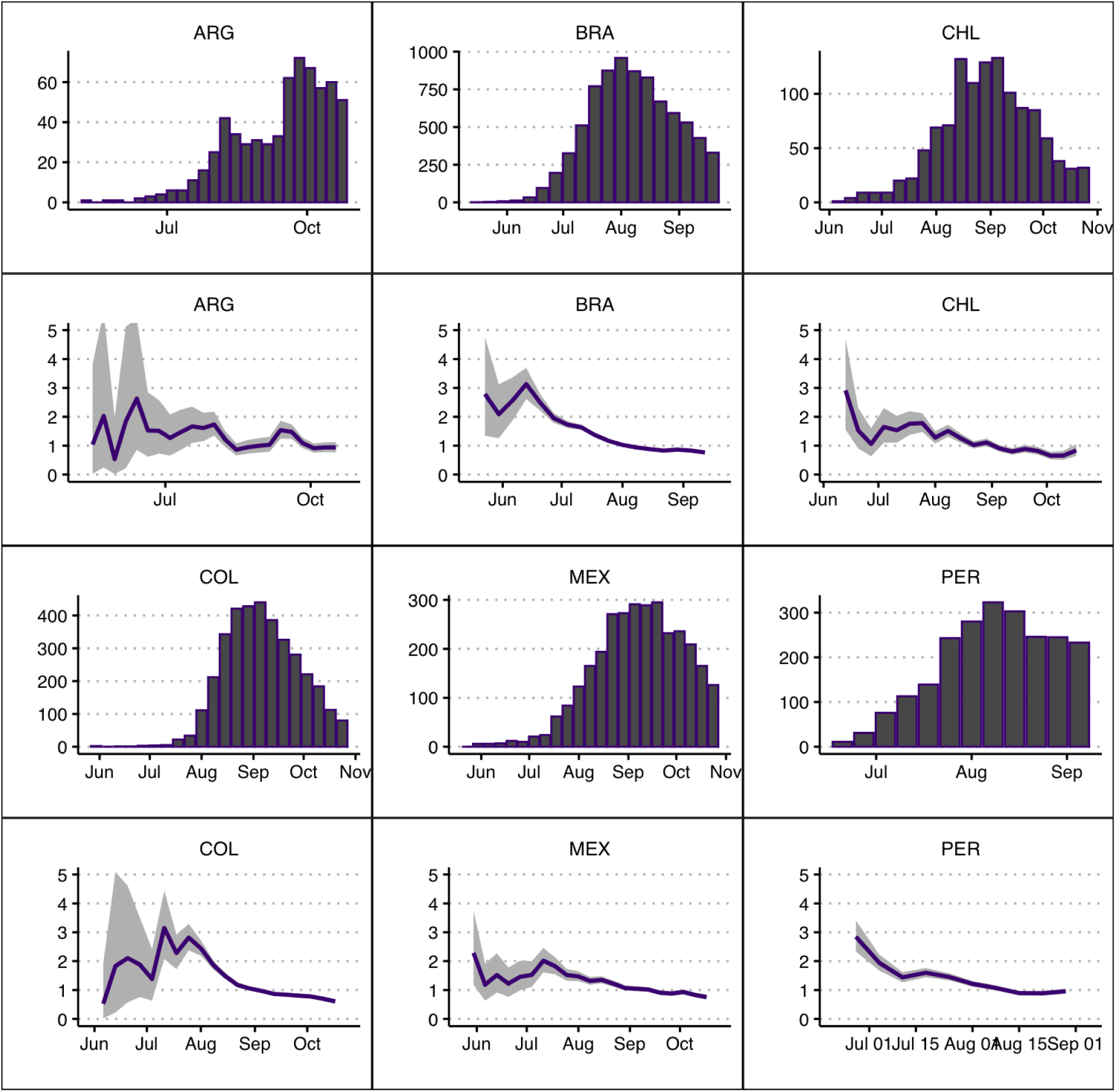
Instantaneous reproductive number R(t) estimations for each country according to date of onset

## Discussion

This work provides relevant estimates of the instantaneous reproductive number R(t) of the Mpox outbreak in Latin America in a comparative manner, in the first six countries with the highest number of confirmed cases in the region. Our results show that the outbreak was at its peak during the period between May and July 2022, reporting R(t) values greater than 1 for all the countries analyzed in this report.

Nevertheless, since mid-August 2022, the epidemic started to decrease in some countries, with current R(t) values less than 1 in all the countries analyzed. It is worth mentioning that using country-level trends may obscure subnational heterogeneity, which probably reflects geographic variation in terms of population density and relevant demographic characteristics and social patterns that could play a role in the dynamics of Mpox transmission (15).

Before the 2022 outbreak, R(t) was estimated to be around 0.08 (95% CI: 0.02 to 0.22) for the West African clade and the Congo Basin clade (16). However, in the current outbreak, early estimations of R(t) values in different countries around the world demonstrated values greater than 1. Kwok et al. (17) estimated a basic reproductive number (Ro) for Mpox as of June 18, 2022, oscillating between 1.50 and 1.70 in England, 1.20 and 1.60 in Portugal and 1.70 and 2.00 in Spain. Later, Du et al. (18) reported R(t) values as of July 22, 2022, ranging between 1.42 and 1.7 in the US, 1.24 and 1.6 in France, 1.17 and 1.2 in Germany, 1.10 and 1.3 in Spain, 1.10 and 1.1 in England and 1.00 and 1.04 in Portugal. Our findings are in line with those obtained by Schrarstzhaupt et al (19), reporting early R(t) estimates that ranged between 1.86 and 2.62 in Federal District of Brasília, 1.54 and 2.00 in Rio de Janeiro, 1.56 and 1.77 in Minas Gerais and 1.48 and 2.16 in São Paulo.

Regarding the implementation of mpox vaccination strategies in Latin America, since August 2022 several countries have initiated procurement processes for Mpox vaccines. However, as of august 2023, only three countries (Chile, Brazil, and Peru) have formally initiated the first stages of their vaccination programs, aimed at priority groups (See **table 3**).

**Table 3:**
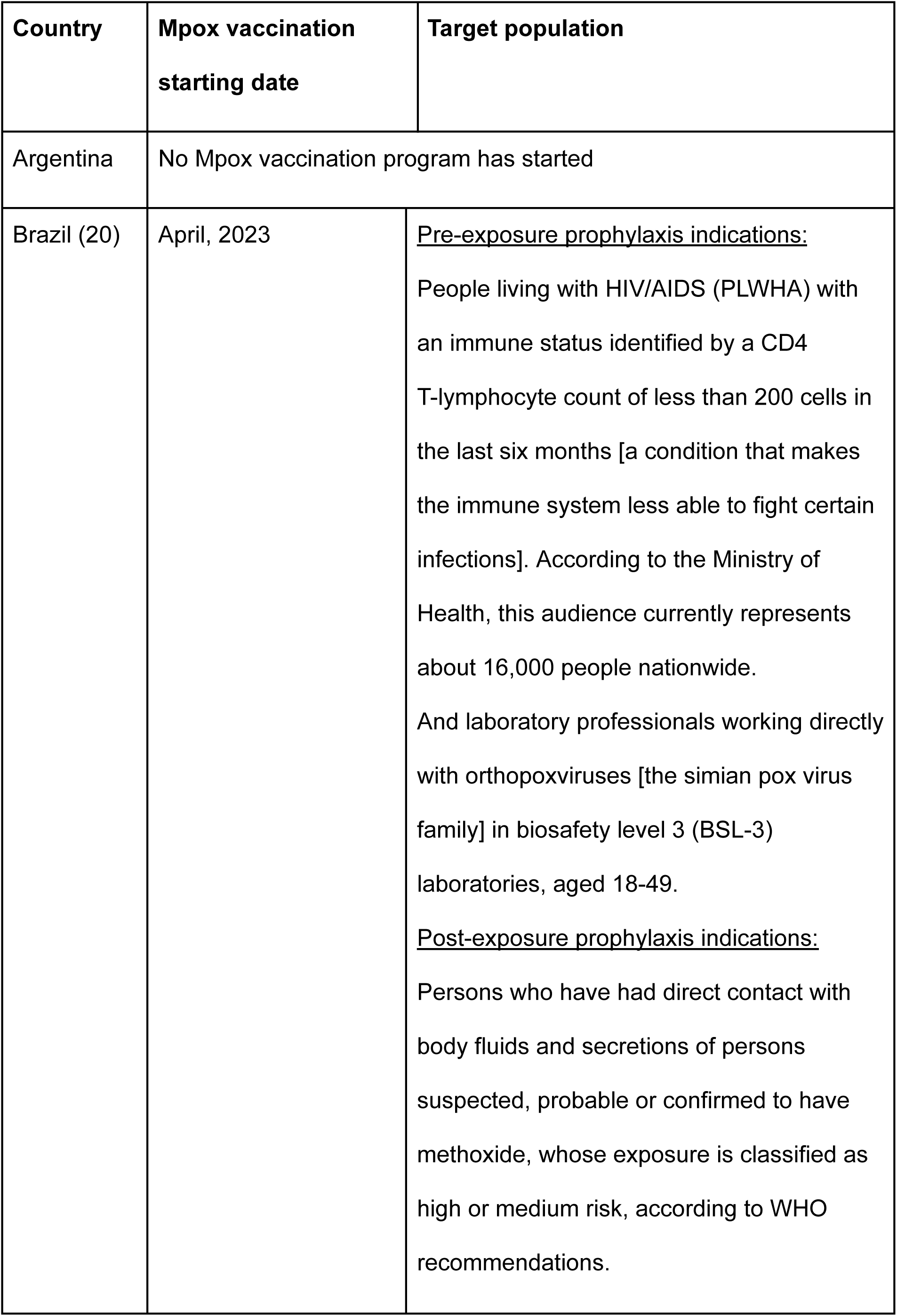
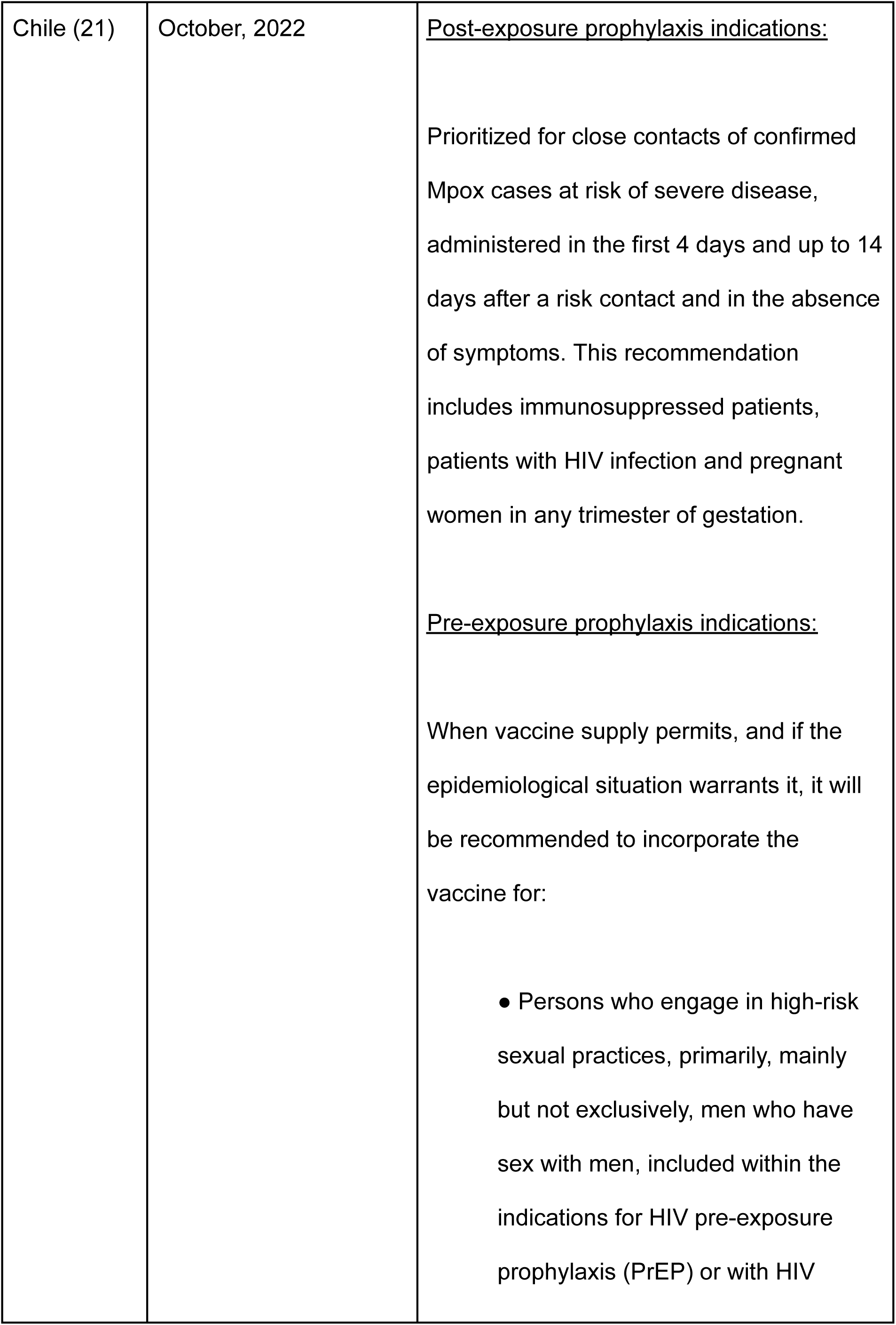
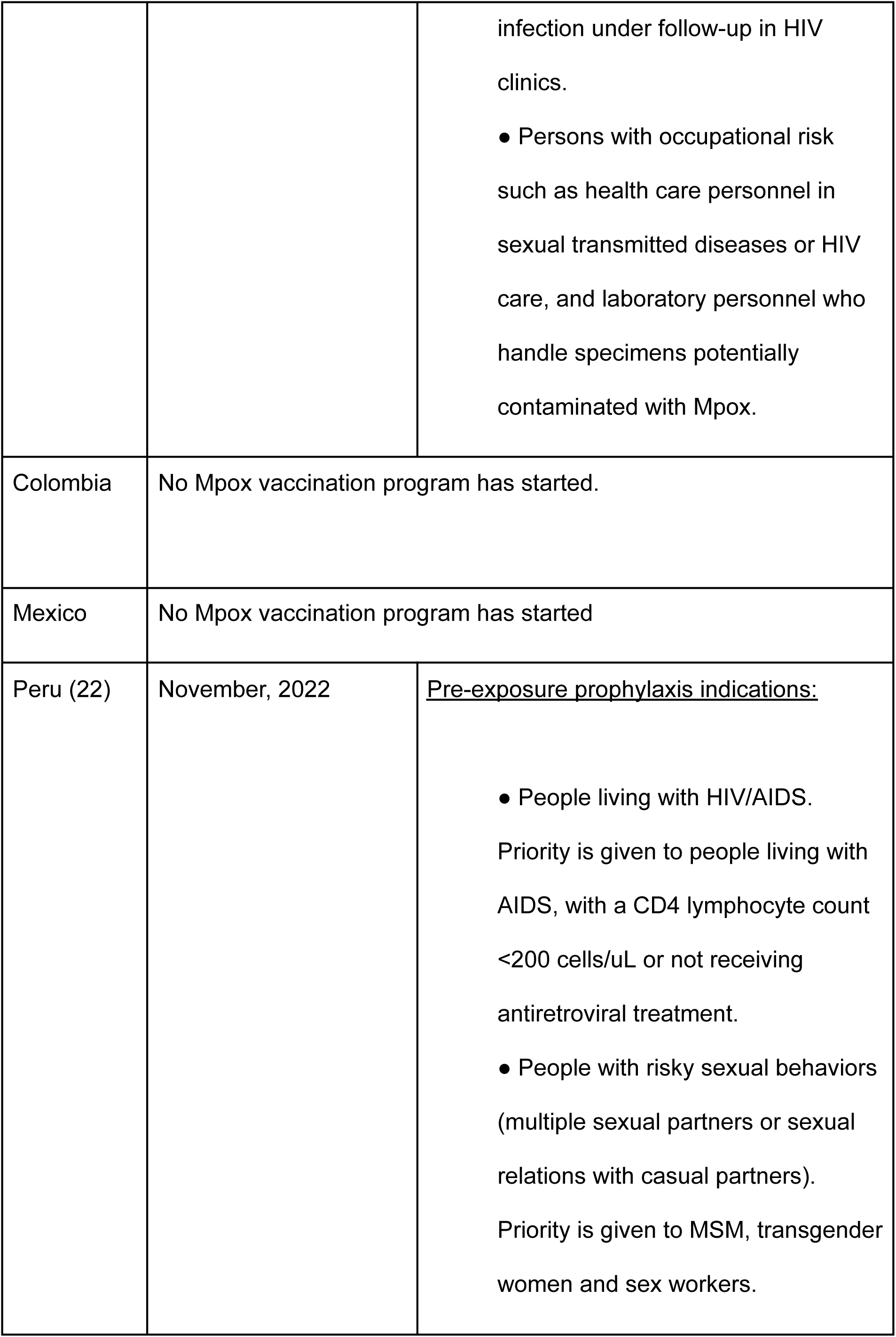
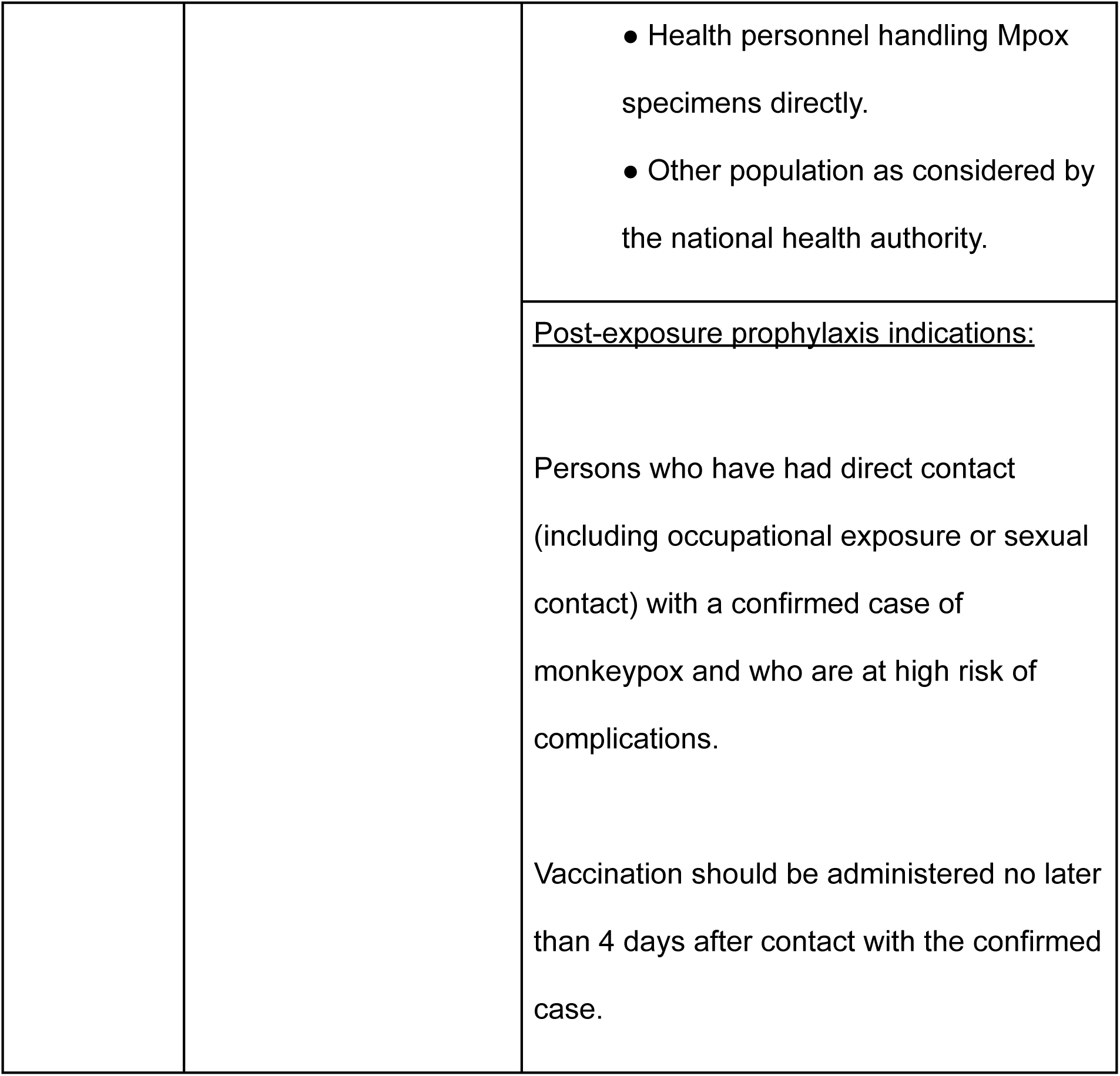
Characteristics of Mpox vaccination programs in some Latin American countries.

While the problem of shortages of vaccines for mpox is global, the vaccination landscape in Latin America is progressing at a slow pace compared to North American and European countries. This situation highlights inequities in access to effective and safe immunization strategies in middle-to low-income countries in the region, which account for a significant percentage of the disease burden.

Our results show that the R(t) is below 1 in Latin America, despite the reduced amount of vaccination programs implemented across the region and shortage of vaccine doses. This can be explained in part by the information presented by health regulatory agencies such as PAHO and the Centers for Disease Control and Prevention (CDC) around the promotion of behavioral changes in the most affected communities (MSM); including measures such as avoiding large social gatherings and reducing the number of casual and anonymous sexual partners (23). Additionally, due to factors such as disparity in access to diagnostic tests, as well as concerns about stigmatization potentially leading to lower frequency of medical consultation for suspected symptoms, the underreporting, and thus the actual attack rate of infection (proportion of people infected during the outbreak), is not yet known with certainty. This information will be essential to elucidate potential future trajectories in Latin America and future health technology needs.

Among the potential limitations of the use of R(t) are the following: 1) Its estimation is based on the assumption of a homogeneous population, in which all individuals have the same probability of having the same number and type of contacts; however, this can be mitigated by making projections for specific populations; 2) R(t) estimations can be unstable during the early stages of the epidemic, because of a limited sample size; therefore, we restricted our analysis to the starting point were R(t) values became more stable; 3) Its calculation is quite sensitive to the value of the serial interval (time elapsed from the onset of symptoms in the primary case to the onset of symptoms in the secondary case), so that errors in the estimation of this parameter can result in a potential source of bias (24).

## Conclusions

Our results provide relevant information about current trends and future scenarios of the Mpox epidemic in Latin America. Despite the latest apparent reduction of the confirmed cases across the globe, a large population remains at risk, and there is a possibility of new waves of the disease as the epidemic continues its course. The strengthening of the surveillance systems around the world would allow us to know the actual attack rate of infection, being key to perform a real-time outbreak analysis. Finally, these findings could be useful for public policy planning in terms of testing, contact tracing and isolation of the confirmed cases at both local and regional levels, highlighting the importance of the unanswered need for the implementation of vaccinations campaigns directed to high-risk groups.

## Ethical responsibilities

### Protection of humans and animals

This research does not use animal nor human material or clinical data from individuals.

### Funding

No direct financial sources were received for the development of the manuscript.

### Contributions of the authors

ZMC wrote the first draft and contributed to writing the results and discussion section of this article. CDB contributed to all the sections in the manuscript. All authors revised and approved the final version.

### Availability of data and materials

The datasets analyzed during the current study are available in the Monkeypox cases - Region of the Americas repository from PAHO and can be sent upon request.

## Data Availability

All data produced in the present study are available upon reasonable request to the authors

## List of Abbreviations

Mpox: monkeypox
WHO: World Health Organization
R(t): instantaneous reproductive number
PAHO: Pan American Health Organization
MSM: male who have sex with male.
CrI: credible interval
SI: serial interval
PrEP: pre-exposure prophylaxis
HIV: Human immunodeficiency virus
AIDS: acquired immunodeficiency syndrome
CDC: Centers for Disease Control and Prevention

